# Sustained High Prevalence of Multiple Antimalarial Drug Resistance Markers in Uganda in 2023-24

**DOI:** 10.64898/2026.03.15.26348231

**Authors:** Thomas Katairo, Victor Asua, Francis D. Semakuba, Bienvenu Nsengimaana, Shahiid Kiyaga, Nicholas Hathaway, Kathryn Murie, Stephen Tukwasibwe, Innocent Wiringilimaana, Jackie Nakasaanya, Caroline Mwubaha, Karen B. Achom, Trevor E. Esilu, Alisen Ayitewala, Monica Mbabazi, Kisakye D. Kabbale, Jerry Mulondo, Eric Watyekele, Andrés Aranda-Diaz, Catherine S. Maiteki, Jimmy Opigo, Grant Dorsey, Samuel L. Nsobya, Moses R. Kamya, Bosco B. Agaba, Isaac Ssewanyana, Bryan Greenhouse, Philip J. Rosenthal, Jessica Briggs, Melissa D. Conrad

**Affiliations:** Infectious Diseases Research Collaboration, Kampala, Uganda; National Malaria Reference and Research Laboratory (NMRRL), Central Public Health Laboratories (CPHL), Kampala, Uganda; Institute of Tropical Medicine, University of Tübingen, Tübingen, Germany; Makerere University, Kampala, Uganda; African Center of Excellence in Bioinformatics and Data Intensive Sciences, Kampala, Uganda; University of California, San Francisco, USA; Uganda Christian University, Mukono, Uganda; Central Public Health Laboratories, Kampala, Uganda; Barcelona Institute for Global Health (ISGlobal), Barcelona, Spain; National Malaria Elimination Division, Kampala, Uganda; Mbarara University of Science and Technology, Mbarara, Uganda; Johns Hopkins Bloomberg School of Public Health, Johns Hopkins University, Baltimore, USA

## Abstract

The emergence and spread of drug resistance threatens malaria control in Uganda. This study reports new data on key polymorphisms associated with antimalarial drug sensitivity at 32 malaria reference centers in Uganda in 2023 and 2024. Samples were collected from patients aged >6 months presenting with uncomplicated falciparum malaria, sequenced using the MAD^4^HatTeR panel and Illumina platforms. Multiple validated or candidate K13 mutations associated with artemisinin partial resistance were detected at ≥20% prevalence at one or more sites across multiple timepoints. The K13 mutations A675V and C469Y predominated in northern Uganda, P441L was most common in western Uganda, and R561H and C469F were confined to southwestern Uganda. After rapid increases in earlier years, prevalences of K13 mutations plateaued at most sites, with A675V, C469Y and P441L reaching maximum site prevalences of 40%, 58%, and 46%, respectively. Prevalence of the chloroquine resistance marker CRT K76T was generally low but increased substantially at three sites in northwestern Uganda, rising to 38% at the last timepoint, with CRT H97L, newly reported in Africa, following a similar trend. The antifolate resistance quintuple mutant haplotype remained highly prevalent nationwide. In addition, DHPS A581G and DHFR I164L, markers of higher-level antifolate resistance, were most common in southwestern Uganda with prevalences up to 64% and 76%, respectively; DHFR I164L also expanded into central and eastern regions. These results highlight continued geographic heterogeneity and underscore the need for continued, nationwide molecular surveillance to guide treatment and chemoprevention policies.

## Introduction

*Plasmodium falciparum* malaria remains a major public health burden in Africa, where nearly 95% of global malaria cases and deaths occur. After gains early this century, progress in malaria control has stagnated over the last decade (WHO 2025). Uganda carries about 5% of the global malaria burden, with an estimated 13.2 million cases in 2024 (WHO 2025). Malaria transmission in Uganda is heterogeneous, with hyperendemic hotspots in the north and east reporting up to 34% prevalence of microscopically detected parasitemia among children under 5 years of age, while the southwest highlands and urban centers have prevalences below 5%, with some areas approaching pre-elimination status (MIS 2019).

Antimalarial drugs are among the most important tools for malaria control. Critical among them are artemisinin-based combination therapies (ACTs), which combine a fast-acting, short-lived artemisinin derivative with a longer-acting partner drug. In Uganda, ACTs replaced chloroquine plus sulfadoxine-pyrimethamine (SP) as the standard treatment for uncomplicated malaria in 2006 due to widespread resistance to those drugs. Artemether-lumefantrine (AL) is the first-line treatment for uncomplicated malaria (Ministry of Health Uganda 2016); the alternative ACTs artesunate-amodiaquine (ASAQ), dihydroartemisinin-piperaquine (DP), and artesunate-pyronaridine (ASPY), are available in private clinics and pharmacies. Severe malaria is treated with intravenous artesunate followed by an oral ACT. For chemoprevention, SP is used alone for malaria prevention in pregnant women and in children with sickle cell anemia, or combined with amodiaquine for seasonal malaria chemoprevention (SMC) in parts of northeastern Uganda (Ministry of Health 2020).

Established and emerging resistance threaten the utility of available antimalarial drugs. Artemisinin partial resistance (ART-R), which first emerged in southeast Asia, manifests as delayed parasite clearance after therapy with an artemisinin, and has been associated with a number of mutations in the Kelch-13 protein (K13) propeller domain (WWARN K13 Genotype-Phenotype Study Group 2019; Ashley et al. 2014; Hamilton et al. 2019). Alarmingly, multiple validated or candidate markers of ART-R, including P441L, C469Y/F, R561H and A675V, have emerged in Uganda, and are now prevalent in different regions of the country (Conrad et al. 2023; Balikagala et al. 2021; Asua et al. 2019, 2021). Prevalences of mutations in MDR1 and CRT that mediate resistance to chloroquine and amodiaquine have decreased since the introduction of ACTs, likely due to selection of wild-type (WT) alleles by lumefantrine and decreased use of aminoquinolines (Asua et al. 2021; P. Tumwebaze et al. 2017; Mbogo et al. 2014). Longitudinal *ex vivo* susceptibility data from northern and eastern Uganda indicate that these changes in parasite genetics have coincided with decreased susceptibility to lumefantrine and dihydroartemisinin, and increased susceptibility to the aminoquinolines chloroquine and amodiaquine (Okitwi et al. 2025; P. K. Tumwebaze et al. 2022, 2021). Furthermore, a recent therapeutic efficacy study in Uganda suggested important shifts in ACT efficacy. In this 2022-2023 study, slow parasite clearance was associated with the presence of K13 mutations, and AL had genotype-corrected efficacy < 90% in northwestern and southeastern Uganda, with efficacy inferior to those of ASAQ, DP, and ASPY in both genotype-uncorrected and -corrected analyses (Kamya et al. 2026).

Resistance to SP, the primary drug used for malaria chemoprevention, is mediated by mutations in the target enzymes dihydrofolate reductase (DHFR) and dihydropteroate synthase (DHPS), the respective targets for pyrimethamine and sulfadoxine. These mutations appear in a stepwise manner and have a cumulative impact on susceptibility (Gregson and Plowe 2005). The quintuple mutation haplotype, comprised of DHFR N51I, C59R, and S108N and DHPS A437G and K540E, is seen at high prevalences across East Africa, including Uganda, and confers an intermediate level of resistance to SP (Conrad and Rosenthal 2019; Gregson and Plowe 2005; Naidoo and Roper 2013). Additional mutations, DHFR I164L, DHPS A581G, and DHPS A613S, mediate higher-level resistance and are selected by IPTp with SP (Kizza et al. 2026; Nayebare et al. 2020; Naidoo and Roper 2013; Harrington et al. 2009). Though historically uncommon in the region, DHFR I164L has been seen at modest prevalences in southwestern Uganda and DHPS A581G in northern Tanzania, eastern Democratic Republic of the Congo and southwestern Uganda (Asua et al. 2019; P. Tumwebaze et al. 2017; Alifrangis et al. 2009; Aydemir et al. 2018).

Here we report new data on key polymorphisms associated with drug sensitivity at 32 sites in Uganda in 2023 and 2024. These data are presented within the context of previously published molecular surveillance studies from 2016-2022 to characterize longitudinal trends across sites with varying malaria epidemiology.

## Results

### Study sites and samples

In 2023-2024, 17,673 samples were collected from symptomatic patients diagnosed with malaria at 32 health facilities across Uganda (**Figure 1**). Of the 10,030 samples selected for sequencing, 8,518 (85%) passed quality control for coverage and stringent checks on sample duplication and were included in downstream analyses (**Table 1**).

**Figure 1.**
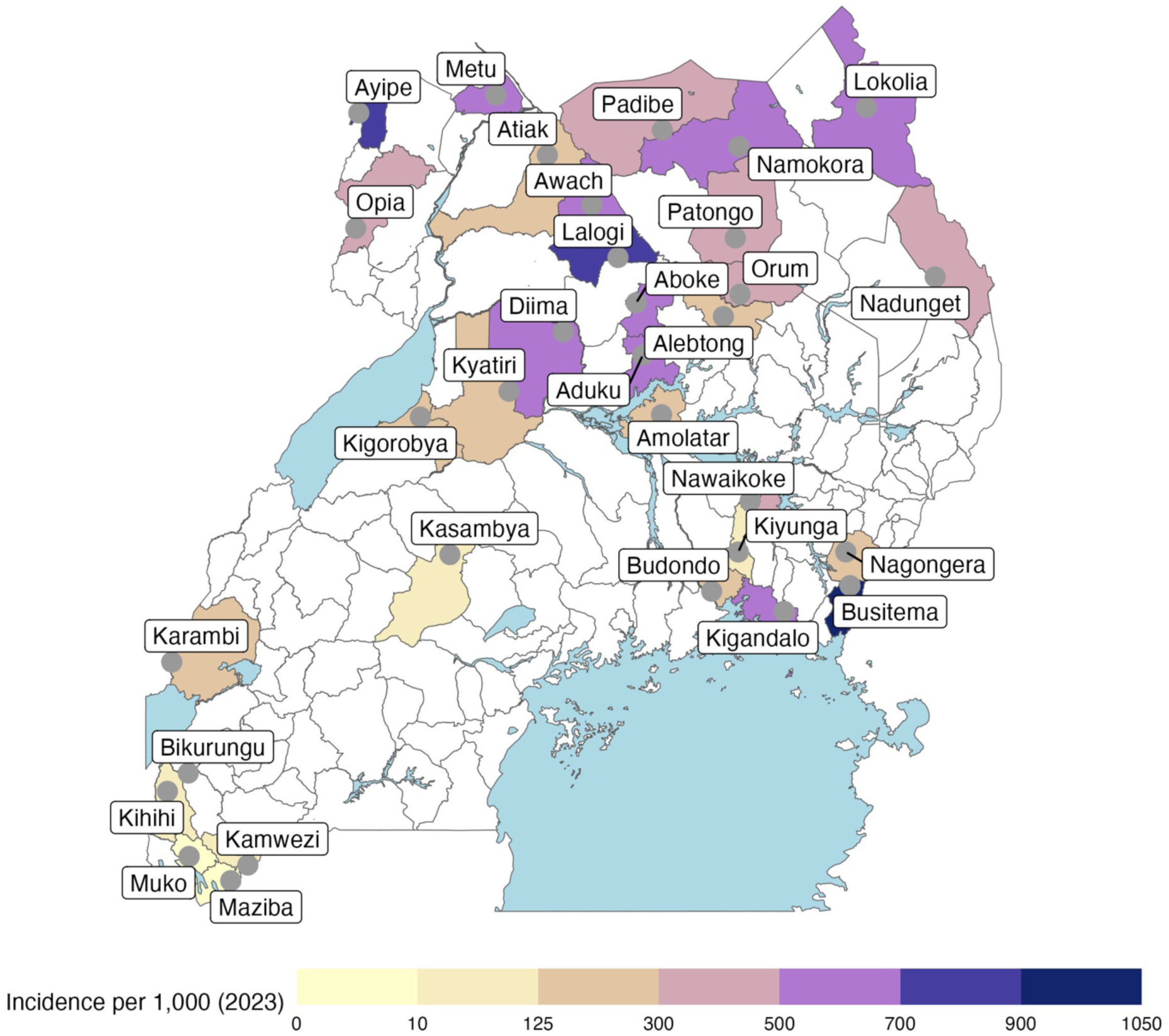
Molecular surveillance sites. Health facilities from which patients were enrolled are indicated by labeled circles. Administrative districts are shaded to represent 2023 malaria incidence per 1,000 person years at the malaria reference center within that district using the provided color scale.

**Table 1.** Number of samples collected, genotyped and analyzed.

| Table 1. Number of samples collected, genotyped and analyzed. |  |  |  |  |  |
| --- | --- | --- | --- | --- | --- |
| Collection | Date of collection | # Sites sampled | # Samples collected | # Samples selected for genotyping | # Samples included in analysis |
| Round 1 | Jan-Jun 2023 | 29 | 4317 | 2513 | 2,365 |
| Round 2 | Jul-Dec 2023 | 28 | 5109 | 2738 | 2,352 |
| Round 3 | Jan-Jun 2024 | 29 | 4368 | 2562 | 2,045 |
| Round 4 | Jul-Dec 2024 | 29 | 3879 | 2217 | 1,756 |
| Total | 2023-2024 | 32 | 17,673 | 10,030 | 8,518 |

The median age of participants providing genotyped samples was < 15 years for all moderate and high transmission sites and ≥ 20 years for low transmission sites (**Supplemental Table 1**). Study participants were more likely to be female at moderate and high transmission sites and male at low transmission sites. Parasite density measured by qPCR varied by site and collection round, with medians of 5,000 - 36,000 parasites/μL (**Supplemental Table 1**).

### Prevalence of *P. falciparum* mutations associated with antimalarial drug resistance

#### Markers of ART-R

Multiple validated/candidate ART-R-mediating K13 mutations were observed at ≥ 20% prevalence at one or more sites in multiple surveillance rounds (**Figure 2**). Two mutations, C469Y and A675V, were observed at high prevalence across northern Uganda and detected at lower prevalence in other regions. The P441L mutation was observed at high prevalence in southwestern and western Uganda. The C469F and R561H mutations were both seen primarily at the Kamwezi malaria reference center (MRC) and at nearby sites in southwestern Uganda.

**Figure 2:**
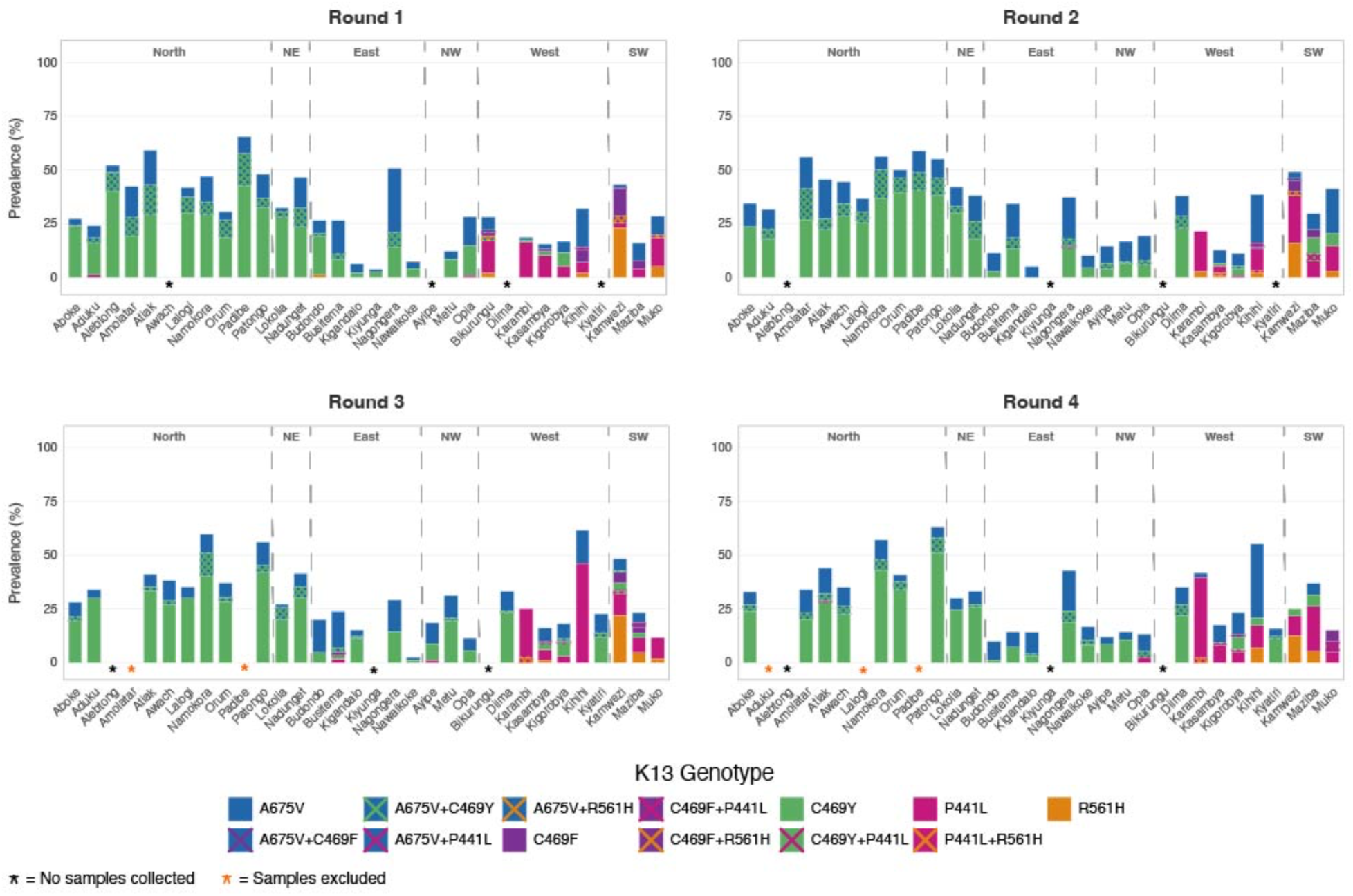
Prevalence of validated/candidate K13 ART-R mutations over 4 rounds of surveillance.

To evaluate the geo-temporal trajectories of K13 mutation prevalence, we considered site-level allele prevalences for the 2023-2024 samples and for previously published data collected in 2016-2022 from 12 sites that had surveillance throughout this period (**Figures 3 and 4**). Most mutations increased in prevalence and expanded geographically over time, while remaining heterogeneous in distribution (**Figure 3, Supplemental Table 2**). However, for most sites, particularly high transmission sites where C469Y and A675V were predominant, the increase in combined prevalence of K13 mutations observed in earlier years slowed, with relatively stable prevalence over recent years at many sites (**Figure 4).**

**Figure 3.**
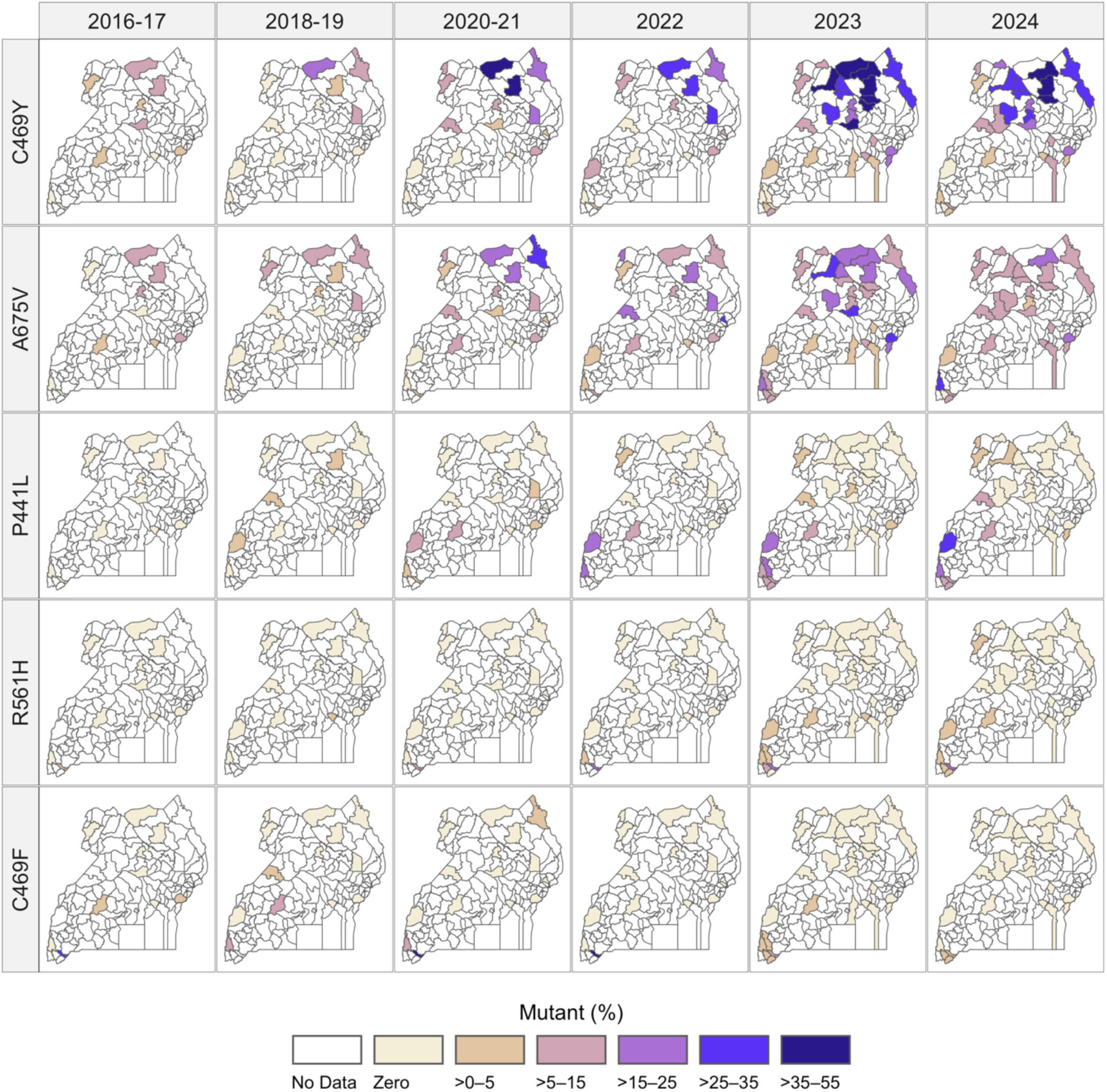
Prevalence of K13 mutations at surveillance sites over time. Administrative districts are shaded to represent prevalence of K13 mutations at the malaria reference center within that district using the provided color scale. Allele frequencies are shown in **Supplemental Figure 1.**

**Figure 4:**
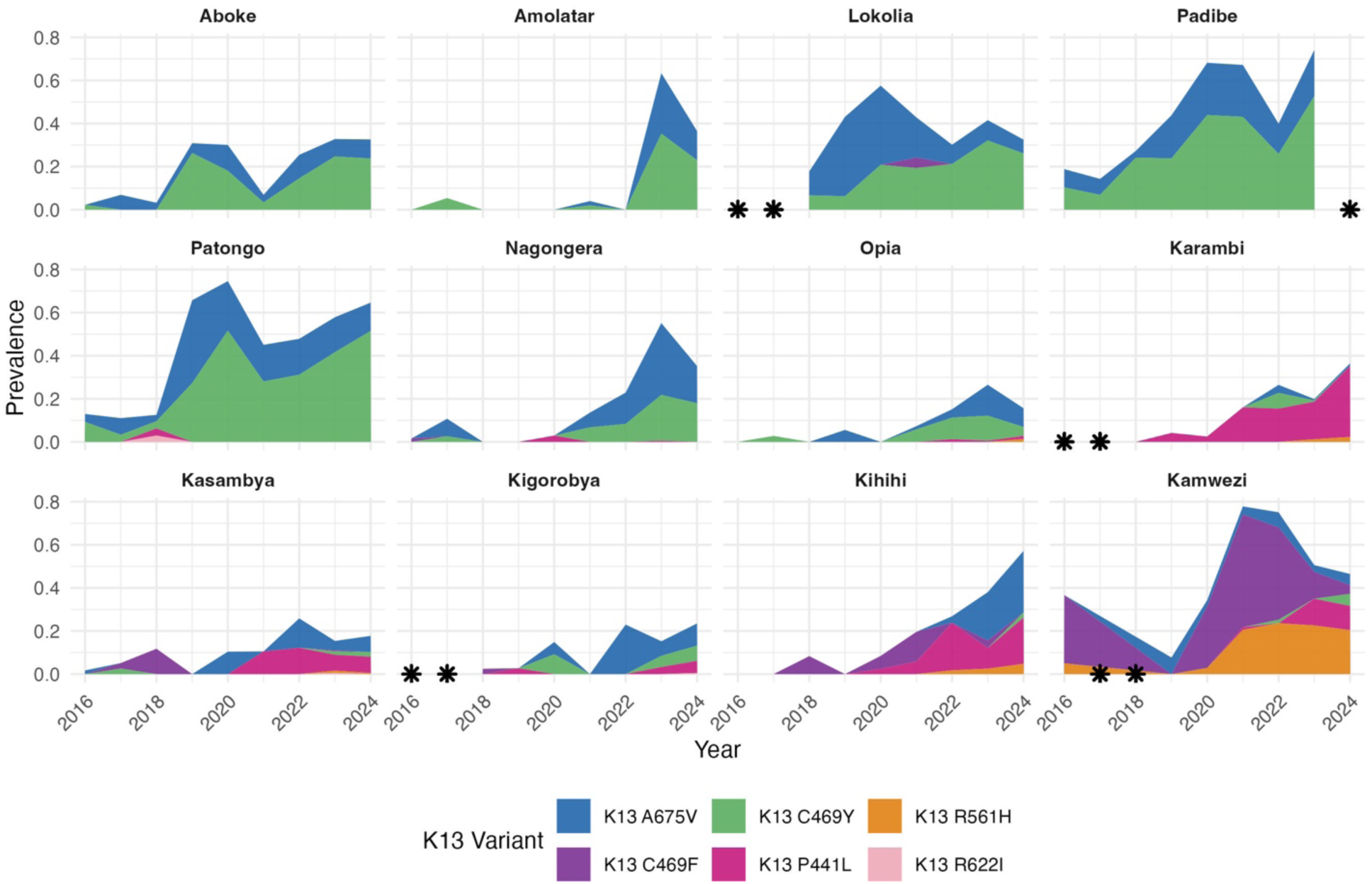
Prevalence of K13 mutations at 12 sites over time. A minimum of 15 genotyped samples was required to estimate yearly prevalences; asterisks indicate years for which prevalence data are not available.

#### Markers of potential ACT partner drug resistance

Prevalence of the CRT K76T mutation was generally low, but in 2023-2024 the prevalence rose in the West Nile region (Ayipe, Metu, and Opia) in Northwestern Uganda, with the mean regional prevalence increasing from 37% in Round 1 to 42% in Round 4 (**Supplemental Figure 2**). In parallel, the CRT H97L mutation, which has been reported at low prevalences in southeast Asia, (Setthaudom et al. 2011; Ye, Zhang, and Zhang 2022) and was previously uncommon in the Uganda (**Figure 5**), reached similar prevalences in West Nile (**Supplemental Figure 1**). Both mutations were seen primarily mixed with WT genotypes, and 99% of samples with the H97L mutation also contained the K76T mutation, suggesting possible genetic linkage. MDR1 N86Y was uncommon in both the historical data and in the 2023-2024 data (**Figure 5, Supplemental Table 2**).

**Figure 5.**
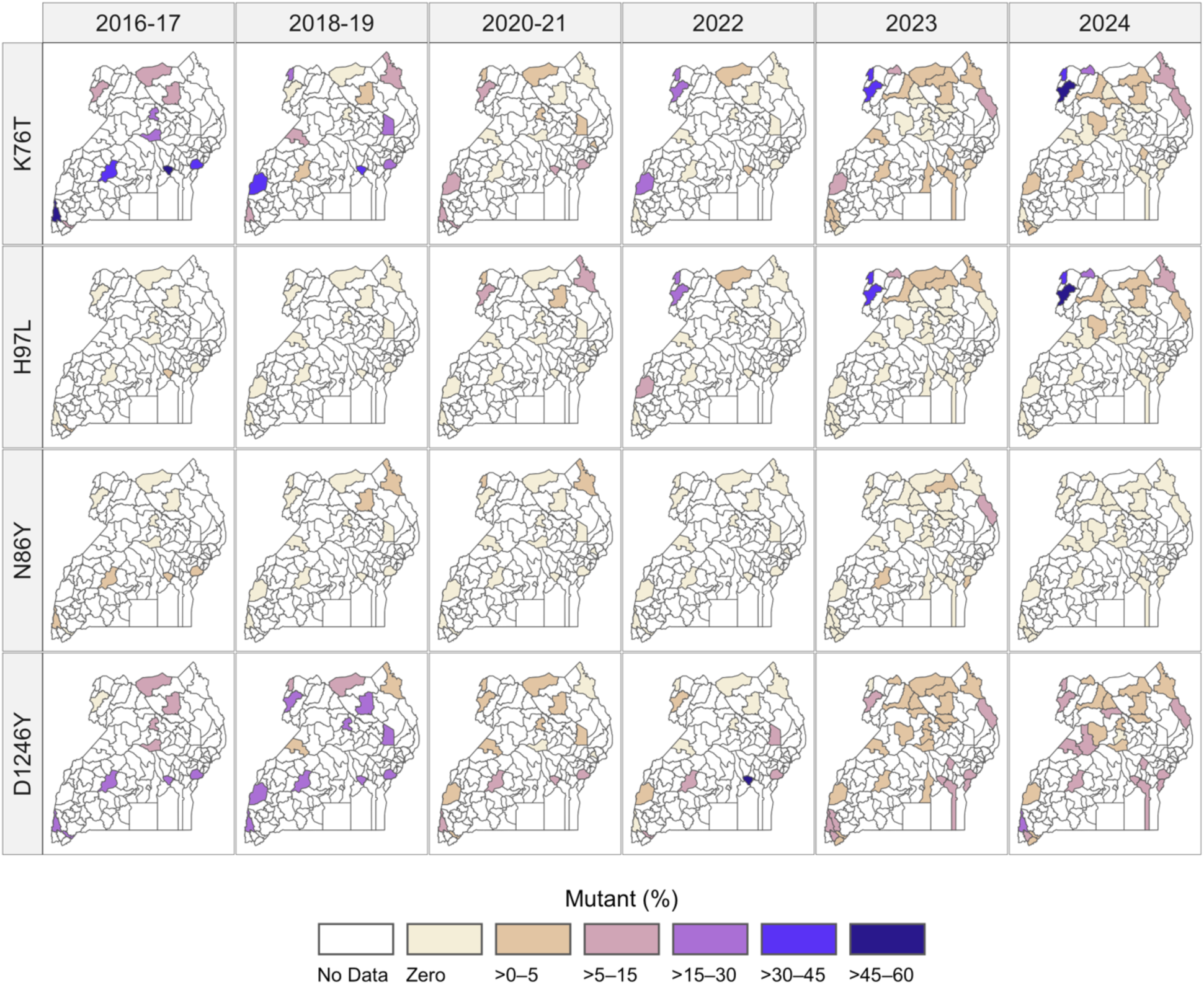
Prevalence of transporter mutations (CRT K76T and H97L, MDR1 N86Y and D1246Y) over time. Shaded districts indicate sampling locations, with color scale representing the prevalence of each mutation at the malaria reference center within that district using the provided color scale. Allele frequencies are shown in Supplemental Figure 3.

We examined copy number variation in *mdr1* and *pm2/3* in 7514 samples from 2023-2024. Over both years, only 0.2% (16/7514) of samples had >1.5 copies of *mdr1* and 0.05% (4/7514) of *pm2/3*, with no clear temporal or geographic pattern (**Supplemental Table 3**).

#### Markers of antifolate resistance

Each of the five mutations that make up the quintuple mutant haplotype (DHFR N51I, C59R, S108N, and DHPS A437G, K540E), which mediates an intermediate level of resistance to SP, was seen at high prevalences across Uganda in 2023-24 (**Supplemental Table 2**). The DHFR I164L and DHPS A581G mutations, which mediate higher-level resistance to SP, were both most common in southwestern Uganda, with DHFR I164L extending to central and eastern regions in recent years (**Supplemental Figure 4**). When phased haplotypes were considered (**Figure 6, Supplemental Table 4**), the quintuple mutation (DHFR N51I, C59R, S108N, I164 and DHPS A437G, K540E, A581, A613) was the most common and widely distributed haplotype (maximum prevalence of 97%), the quintuple + I164L haplotype was prevalent across southern Uganda (maximum of 55%), and the quintuple + A581G and the septuple mutation (DHFR N51I, C59R, S108N, I164L and DHPS A437G, K540E, A581G, A613) haplotypes were prevalent in southwestern Uganda (maximum of 54% and 37%, respectively).

**Figure 6.**
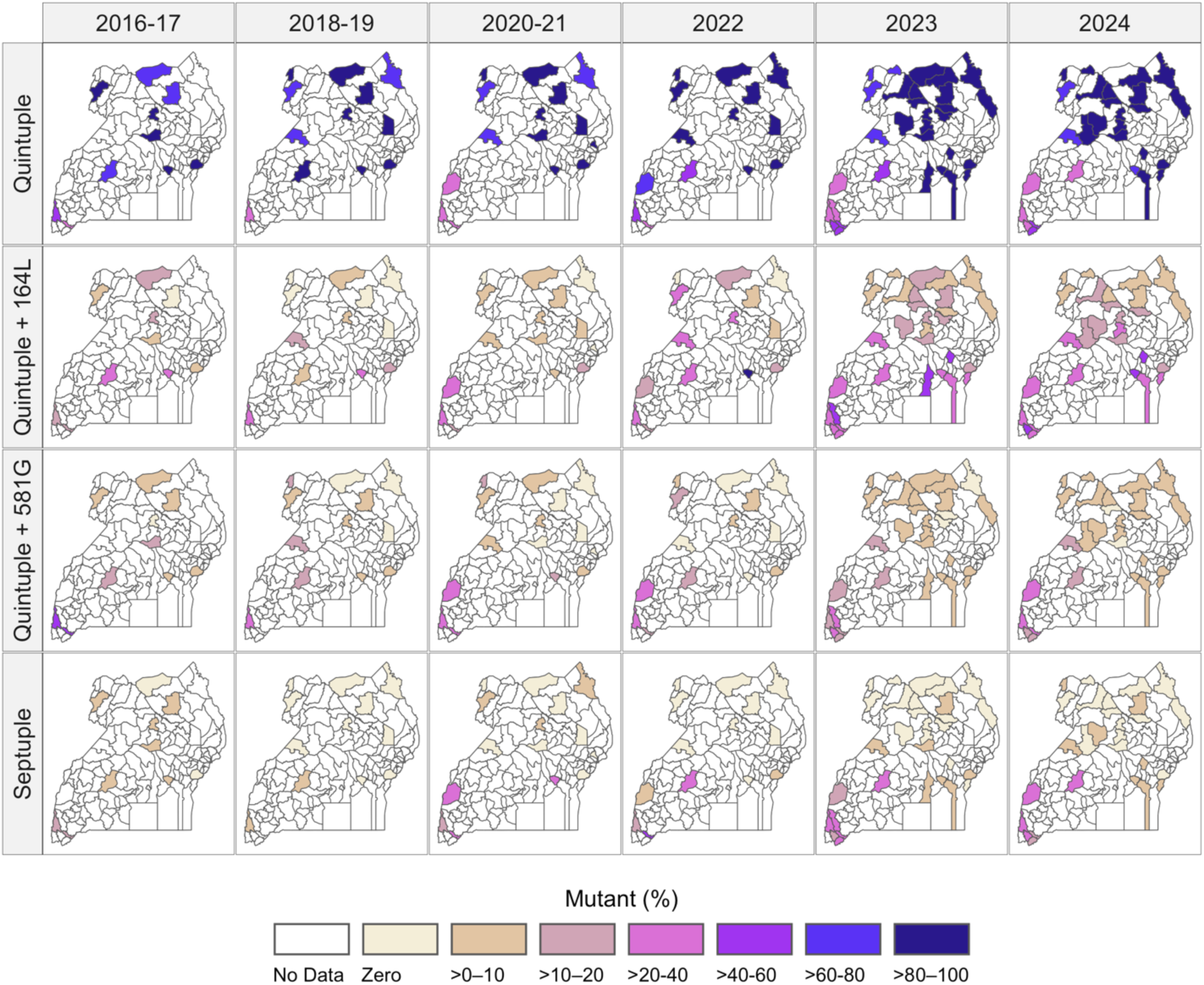
Prevalence of antifolate resistance haplotypes (DHFR N51I, C59R, S108N, I164L and DHPS A437G, K540E, A581G). The quintuple haplotype is defined as **IRN**I**GE**A, quintuple + 164L as **IRNLGE**A, quintuple + 581G as **IRN**I**GEG**, and the septuple haplotype as **IRNLGEG**. Shaded districts indicate sampling locations, with color scale representing the prevalence of each mutation at the malaria reference center within that district using the provided color scale. Haplotype frequencies are shown in Supplemental Figure 5.

## Discussion

The emergence of markers of artemisinin partial resistance in *P. falciparum* populations, evidence of decreasing susceptibility to lumefantrine (Okitwi et al. 2025; P. K. Tumwebaze et al. 2022, 2021), and suboptimal performance of AL in therapeutic efficacy trials (Kamya et al. 2026) all raise fears that AL, the most widely used ACT in sub-Saharan Africa, is beginning to fail. Also concerning is the increasing prevalence of *P. falciparum* markers of resistance to SP, raising questions about the utility of this drug for chemoprevention. We report prevalences and frequencies of key markers of antimalarial drug resistance from isolates causing symptomatic malaria infections at 32 MRCs across Uganda in 2023-2024. Our data show sustained high prevalence and marked geographic variation of markers of resistance to artemisinins and antifolates, regional emergence of mutations associated with aminoquinoline resistance, and near fixation of a marker of decreased susceptibility to lumefantrine.

Combined prevalences of five validated/candidate K13 mutations associated with ART-R were high across Uganda, surpassing 25% at many sites. Consistent with previous reports (Conrad et al. 2023), the distribution of the five mutations was geographically heterogeneous, with the C469Y and A675V mutations widely distributed across the north and east of the country, the P441L mutation primarily located along the western border, and the C469F and R561H mutations generally limited to relatively low-transmission sites in the Southwest. However, in contrast to modeled data suggesting that K13 mutations were moving toward fixation (Meier-Scherling et al. 2025; Nguyen et al. 2025), these newer data indicate that mutation prevalence has plateaued at many sites. These results suggest there may be a balance between the advantages of relative drug resistance and some loss of fitness for these key mutations, and that this balance may limit the continued rise of ART-R in *P. falciparum* in Uganda.

The selective forces that have resulted in the observed population structure for the K13 locus are unclear. While most of southwestern Uganda has relatively low transmission relative to the rest of the country, Kamwezi is epidemic prone, oscillating between low and moderate transmission (Epstein et al. 2026). The MRCs in northern, western and eastern Uganda are medium to high transmission sites, with varied histories of vector control interventions. Western and northern Uganda may also be influenced by treatment policies in neighboring DRC and South Sudan, where ASAQ has been used as a first-line ACT(WHO 2025) and where self-medication is common (Akilimali et al. 2022; Kayiba et al. 2023).

After the rapid decline of the CRT K76T mutation, the main marker of resistance to CQ, over the first two decades of this century, the subsequent emergence of the CRT K76T and H97L mutations in northwestern Uganda has likely been driven by the use of ASAQ in nearby regions of South Sudan (where ASAQ is the first-line regimen for uncomplicated malaria) and DRC (where ASAQ is one of the first-line regimens). Interestingly, prevalence of the CRT H97L mutation, which was not previously described in Africa, has increased in northwestern Uganda in concert with increased prevalence of K76T, suggesting genetic linkage. Of note, another mutation at the same locus, H97Y, has been associated with decreased DP efficacy in southeast Asia (Hamilton et al. 2019), but not Africa; thus characterizing the phenotype associated with this emerging haplotype is a high priority.

The DHFR/DHPS quintuple mutant continues to be widespread across Uganda, and two additional mutations, DHFR I164L and DHPS A581G, have reached high prevalences, especially in southwestern Uganda. In a recent trial in eastern Uganda, the 28-day cumulative risk of recrudescence after IPTp-SP was 40%, with selection for I164L and A581G after SP exposure (Kizza et al. 2026). These findings, together with low chemopreventive efficacy of SP observed in prior trials in children (Nankabirwa et al. 2010) and pregnant women (Kakuru et al. 2025), raise serious questions about the continued utility of SP-based chemoprevention in Uganda. Of particular concern is the recent implementation of SMC with SPAQ in the Karamoja region: although a cluster randomized trial reported high protective efficacy of both SPAQ and DP (Nuwa et al. 2025), a pharmacometric assessment in the same region found substantially higher rates of parasitemia and symptomatic malaria after SPAQ than DP, with data suggesting that the preventive efficacy of SPAQ was driven primarily by AQ rather than SP (Bonnington et al. 2025). Given that I164L and A581G prevalences are considerably higher in other regions of Uganda relative to Karamoja, SPAQ may offer limited benefit over amodiaquine alone in much of the country while risking further selection of amodiaquine resistance. These findings underscore the urgent need to reconsider SP-based chemoprevention strategies in Uganda and other settings in eastern Africa where high-level SP resistance is well established.

Our data demonstrate dynamic changes in prevalences of multiple *P. falciparum* drug resistance markers in Uganda over time. Key trends include (a) increasing prevalence of multiple K13 mutations that mediate ART-R, with stabilization of prevalence at many sites; (b) reappearance of the CRT K76T mutation, accompanied by the newly detected H97L mutation, in northwestern Uganda, suggesting a recurrence of resistance to chloroquine and amodiaquine in that region; and (c) high and increasing prevalence of markers of high level resistance to SP, consistent with increasing resistance to this drug. These data indicate the need for continued surveillance for antimalarial drug resistance, including molecular surveillance as described in this report, measures of ex vivo antimalarial drug activity (Tumwebaze 2022; Okitwi 2025), and clinical trials comparing the antimalarial efficacies of leading regimens (Kamya et al. 2026). Further, we should urgently consider changing policy for treatment and chemoprevention to better confront the spread of resistance, including utilization of multiple first-line regimens, rotating regimens, new combinations including triple ACTs, and new drugs expected to be approved soon.

## Materials and methods

### Collection of samples (2023-2024)

Dried blood spot (DBS) samples were obtained by convenience sampling from symptomatic individuals > 6 months of age presenting with uncomplicated, microscopically confirmed falciparum malaria using thick smears made by pre-trained microscopists and stained by Giemsa at 32 MRCs across Uganda **(Figure 1, Supplemental Table 1)**. For moderate to high transmission MRCs, ∼200 samples were collected biannually (Collection 1: Jan–April 2023, Collection 2: July–Sep 2023, Collection 3: Feb–April 2024, and Collection 4: Aug–Oct 2024). Collections at certain sites were dropped or added through the course of the study for logistical reasons, as follows: Kiyunga, Alebtong, and Bikurungu were dropped after Collection 1; Ayipe, Diima, and Awach were added after Collection 1; and Kyatiri was added after Collection 2. For low-transmission or epidemic-prone MRCs, year-round collections were performed (**Supplemental Table 1**). Biannual and longitudinal samples were grouped by timing of collection, as follows: Round 1: Jan-Jun 2023, Round 2: July-Dec 2023, Round 3: Jan-Jun 2024, Round 4: July-Dec 2024.

### DNA extraction and quantification

DNA was extracted from DBS using Chelex-100/Tween-20, as previously described (Katairo et al. 2025). *P. falciparum* DNA was quantified using *var*ATS-based qPCR, following established methods (Hofmann et al. 2015).

### Library preparation and sequencing

To ensure efficient genotyping, qPCR-quantified samples with parasite densities ≥ 1000 parasites/microliter (µL) were prioritized for sequencing. For sites with > 100 available samples with parasite densities ≥ 1000 parasites/µL, 100 samples were randomly selected for sequencing. For sites with < 100 samples with parasite densities ≥1000 parasites/µL, all samples meeting this threshold were included, with additional samples selected in descending order of parasite density to achieve a total of 100 samples. For low-transmission sites, all samples with parasite densities ≥ 10 parasites/µL were sequenced. Library preparation was conducted using primer pools D1.1, R1.2 and R2.1 of the Multiplex Amplicons for Drug, Diagnostic, Diversity, and Differentiation Haplotypes using Targeted Resequencing (MAD^4^HatTeR) protocol, as previously described (Aranda-Díaz et al. 2025; Katairo et al. 2025). MAD^4^HatTeR targets 165 highly diverse microhaplotypes for genetic diversity and relatedness assessment and 118 key markers for drug and diagnostic resistance (including duplications and deletions). Sequencing was performed at the Central Public Health Laboratory in Uganda using the Illumina MiSeq platform for Rounds 1–3 and both MiSeq and NextSeq platforms for Round 4. Raw sequencing reads have been deposited in the NCBI Sequence Read Archive (SRA) under BioProject accession number PRJNA1429107.

### Quality control

Coverage metrics and allele calls were generated using the MAD^4^HatTeR bioinformatic pipeline v0.2.2 (Aranda-Díaz et al. 2024, https://github.com/EPPIcenter/mad4hatter). To identify potential issues with sample collection and library preparation, sample similarity was evaluated by calculating the Jaccard index and root mean square error of WSAF for highly diverse targets (pool D1.1). Nearly identical WSAF distributions in polyclonal infections, defined as Jaccard index ≥0.70 and root mean square error <0.10, indicative of sample duplication prompted exclusion of data. All data from a collection were excluded when >20% of samples collected from a site were suspected to be duplicated (Amolatar and Padibe in Round 3 and Aduku, Lalogi, and Padibe in Round 4 (**Supplemental Table 1**)). For collections where suspected sample duplication occurred at lower prevalence, a single sample was retained from each duplicated group.

### Presentation of historical data (2016-2022)

To contextualize the 2023-2024 drug resistance data and more clearly demonstrate longitudinal trends, data for samples collected from 2016-2022 were incorporated into our analyses. Sample collection, genotyping methods (using molecular inversion probe (MIP) captures) and prevalence data have been reported previously (Conrad et al. 2023; Asua et al. 2021, 2019). Briefly, 50 samples (2016-2019) and 100 samples (2020-2022) were collected by convenience sampling at 10 sites in 2016 and 2017 and 16 sites from 2018 to 2022 from symptomatic patients > 6 months of age who presented with uncomplicated falciparum malaria at MRCs. Raw sequencing reads were deposited in the NCBI Sequence Read Archive (SRA) under BioProject accession numbers PRJNA880926 (2016 samples), PRJNA880930 (2017 samples), PRJNA655702 (2018 and 2019 samples), PRJNA880932 (2020 samples), and PRJNA880933 (2021 samples) at time of publication.

### SNP calling and estimating prevalence and frequency of drug resistance mutations

The *plasmodiumdrugres* pipeline was used to generate amino acid calls and to estimate the prevalence and allele frequency of single- and multilocus drug resistance mutations within the population (https://github.com/PlasmoGenEpi/plasmodiumdrugres). Genotypes were filtered on a per-sample basis using the following criteria: (i) total read coverage at the target ≥ 50 reads; (ii) ≥ 10 reads supporting the specific allele; and (iii) within-sample allele frequency (WSAF) ≥ 0.75%. Estimates were calculated per site and collection round for MAD^4^HatTeR data, and per site and year for combined MAD^4^HatTeR and MIP data.

Single-locus prevalence was defined as the proportion of samples carrying a given mutation, with both mixed and fully mutant genotypes classified as mutant. Single-locus allele frequency (i.e., the proportion of parasites in the population carrying a mutation) was calculated by weighting alleles according to their within-sample allele frequency (WSAF), derived from read counts for MAD^4^HatTeR data and from unique molecular identifiers (UMIs) for MIP data.

Multilocus estimates were generated for each predefined haplotype using a two-step, model-free approach. First, samples in which only one locus within the grouping had multiple amino acid calls were identified. For these samples, haplotypes were unambiguously inferred by pairing the single amino acid calls at fixed loci with each variant observed at the single variable locus. The WSAF of each inferred haplotype was assigned as the WSAF of the corresponding variant at the variable locus. Samples not meeting this criterion were processed by filtering all loci within the grouping at WSAF ≥ 0.7. If a single amino acid call remained at every locus after filtering, the resulting multilocus allele was assumed to be present. Samples resolved by both approaches were then pooled. Multilocus prevalence was calculated as the proportion of samples carrying each multilocus allele (with mixed and fully mutant genotypes classified as mutant), and multilocus allele frequency was calculated by weighting haplotypes according to their WSAF.

Downstream data analysis was conducted using R version 4.2.2.

### Copy number analysis

For determination of *plasmepsin 2/3* and *mdr1* gene copy number, we applied a generalized additive model to normalize read depth and estimate fold change across target amplicons in the MAD^4^HatTeR data for the *plasmepsin2* (*pm2*), *plasmepsin3* (*pm3*), and *mdr1* genes as described previously (Aranda-Díaz et al. 2025). Because duplications in *pm2* and *pm3* nearly always occur together as a single structural event (Amato et al. 2017), we calculated a single combined fold change across both genes for *plasmepsin* copy number. A sample was considered positive for duplication if the estimated fold change was ≥ 1.5x.

### Ethical considerations

Ethical approval was obtained from the Makerere University School of Biomedical Sciences Research Ethics Committee (protocols SBS-2021-167 and SBS-2024 -566), the Uganda National Council for Science and Technology (protocols HS2309ES and HS5584ES), and the University of California, San Francisco Institutional Review Board (reference 341609). Written informed consent was obtained from all adult participants, with additional assent from children aged 8–18 years.

## Acknowledgements

The study was funded by the Gates Foundation (INV-035751, INV-037316) and the National Institutes of Health (R01AI173557, U19AI089674, K24AI144048, K23AI166009). The funders had no role in study design, data collection and interpretation, or the decision to submit the work for publication. We thank Prof. David P. Katete, Dr. Daudi Jjingo and Prof. Moses Joloba for their extensive support during the implementation of the IMMRSE-U study. We also thank Kylie Hilton for developing data management tools.

## Data Availability

Sequence data that support the findings of this study have been deposited in the NIH NLM Sequence Read Archive (SRA) with the primary accession code PRJNA1429107. Additional data and code are available at 10.5281/zenodo.18854846.

