## Supplemental Figures and Tables for "Sustained High Prevalence of Multiple Antimalarial Drug Resistance Markers in Uganda in 2023-24"

**Supplemental Figure 1: Frequency of K13 mutations at surveillance sites over time.** Administrative districts are shaded to represent frequency of K13 mutations at the malaria reference centre within that district using the provided colour scale.

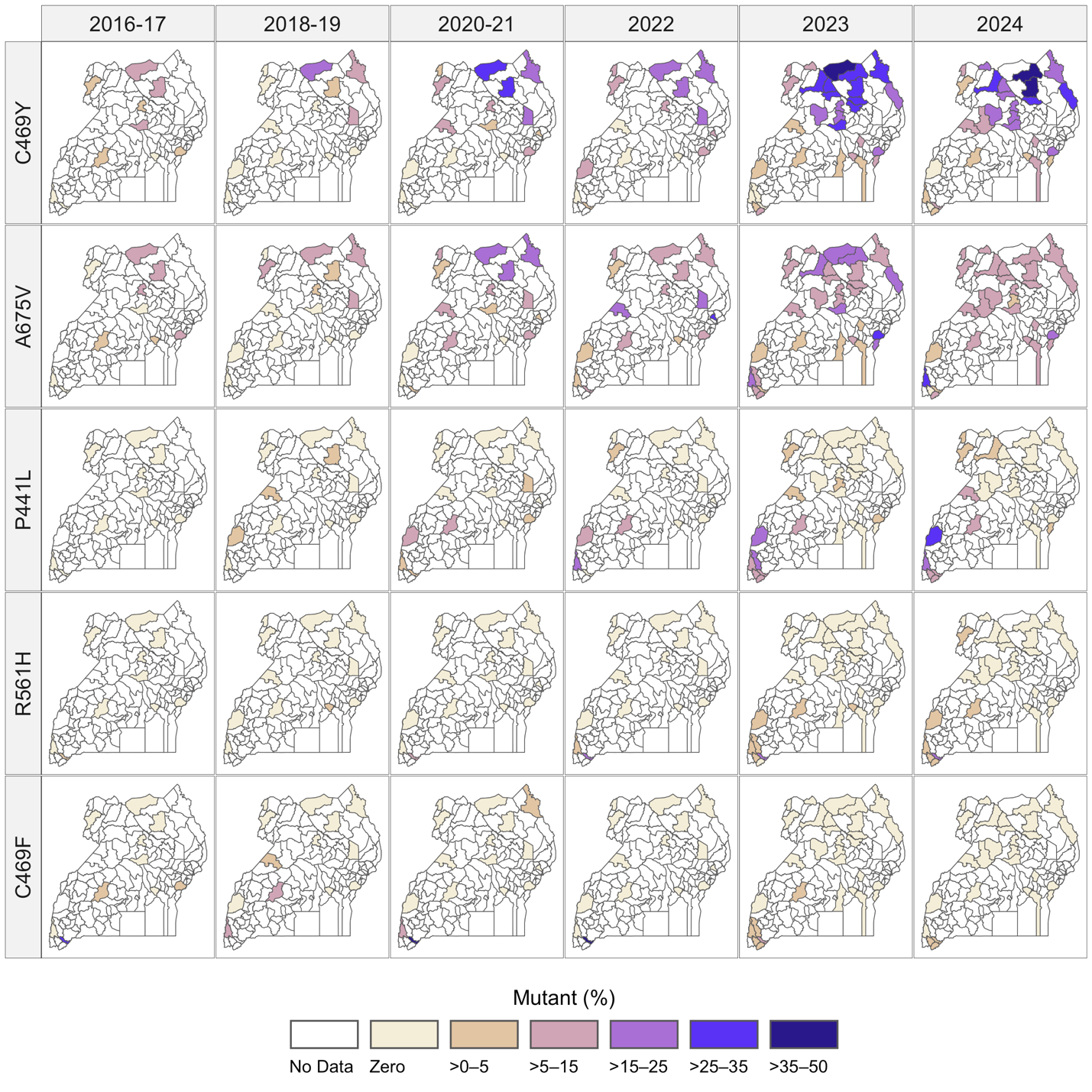

**Supplemental Figure 2: Prevalence of the CRT K76T and CRT H97L mutations over four rounds of surveillance**

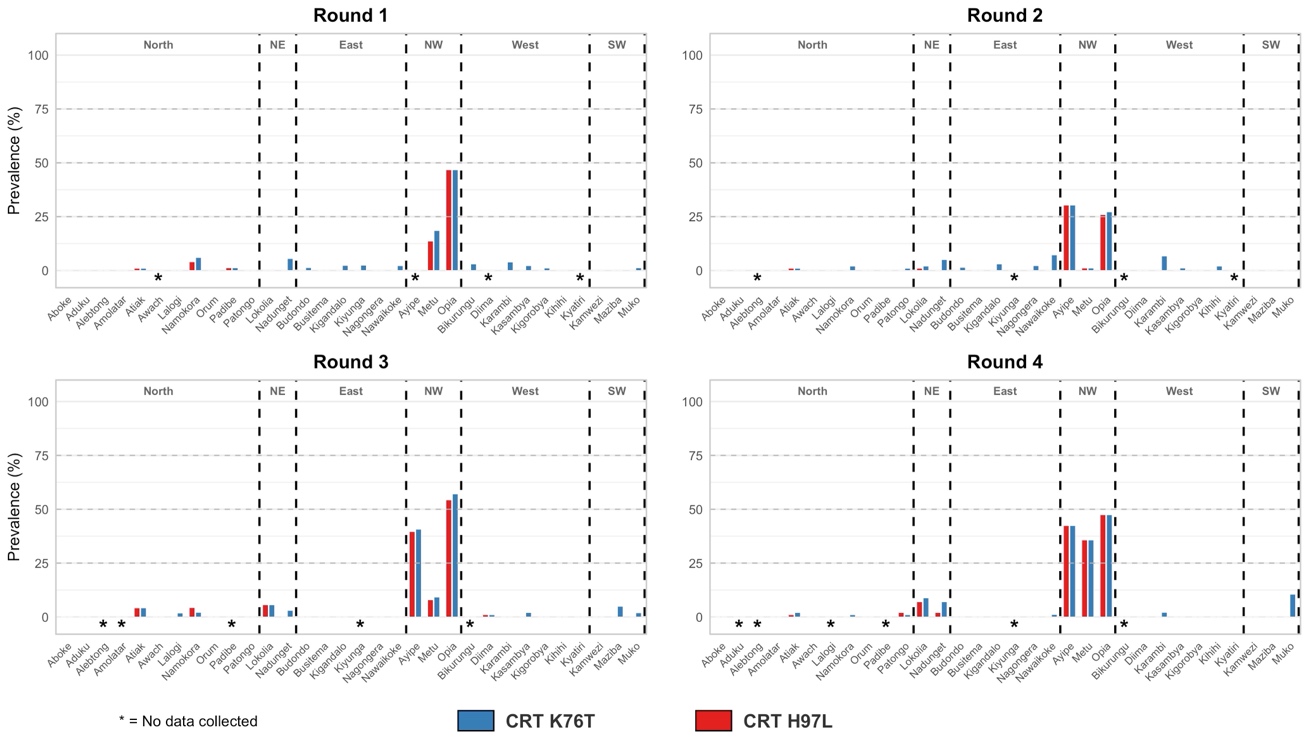

**Supplemental Figure 3. Frequency of transporter mutations (CRT K76T and H97L, MDR1 N86Y and D1246Y) over time.** Shaded districts indicate sampling locations, with colour scale representing the frequency of each mutation at the malaria reference centre within that district using the provided colour scale.

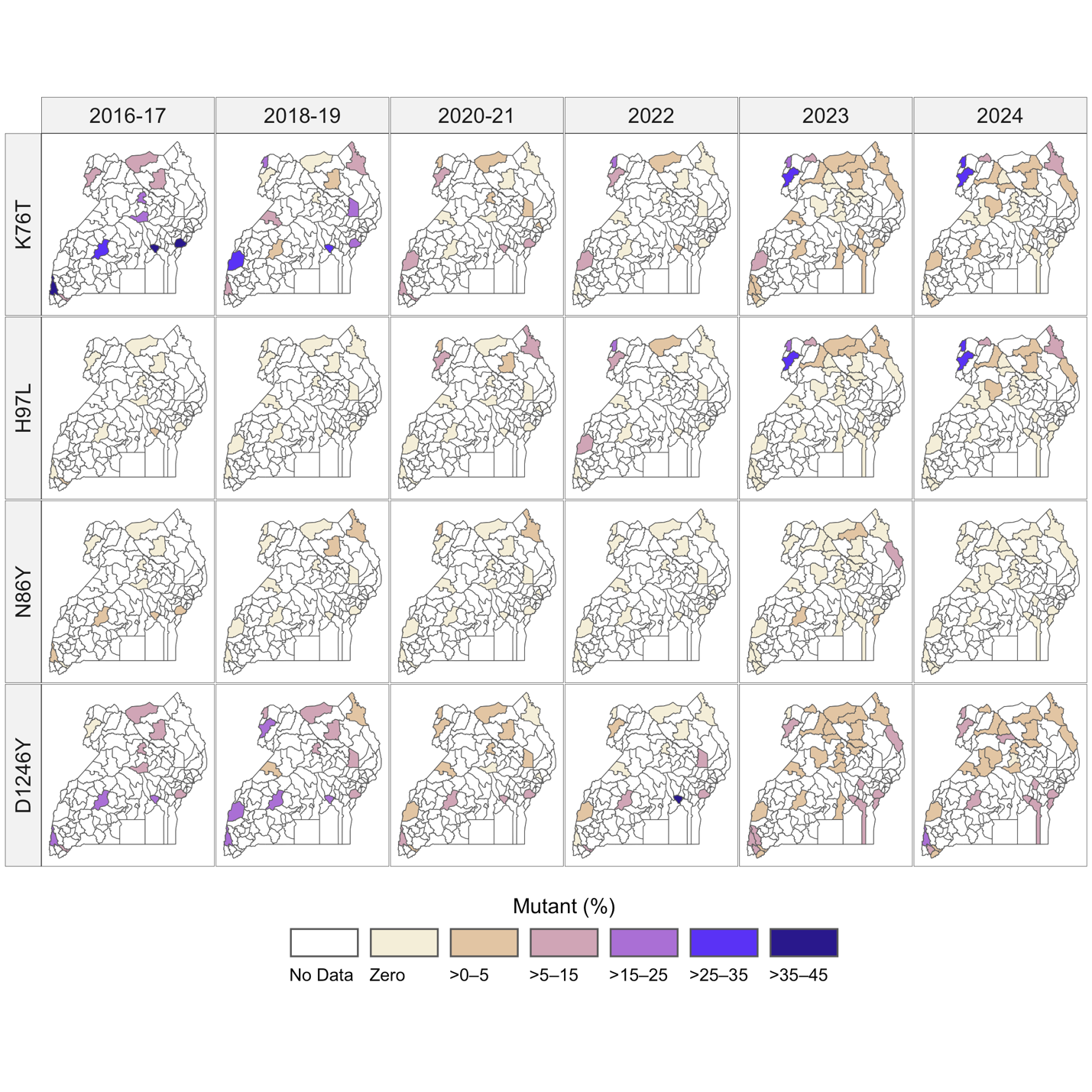

**Supplemental Figure 4: Prevalence of the DHFR I164L and DHPS A581G mutations across the four rounds of collection.**

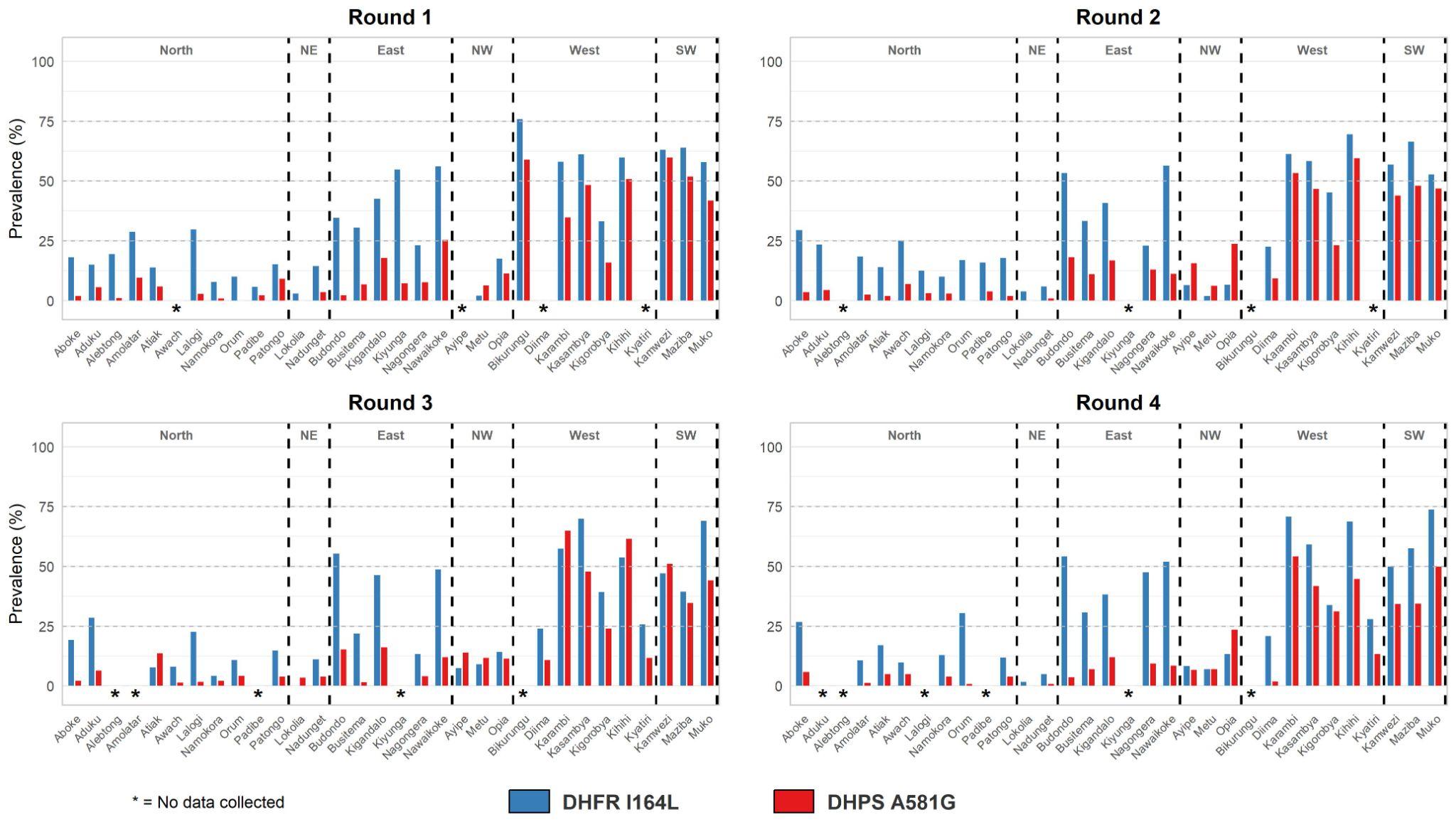

**Supplemental Figure 5. Frequency of antifolate resistance haplotypes (DHFR N51I, C59R, S108N, I164L and DHPS A437G, K540E, A581G).** The quintuple haplotype is defined as **IRN**I**GE**A, quintuple + 164L as **IRNLGE**A, quintuple + 581G as **IRN**I**GEG**, and the septuple haplotype as **IRNLGEG**. Shaded districts indicate sampling locations, with colour scale representing the frequency of each mutation at the malaria reference centre within that district using the provided colour scale.
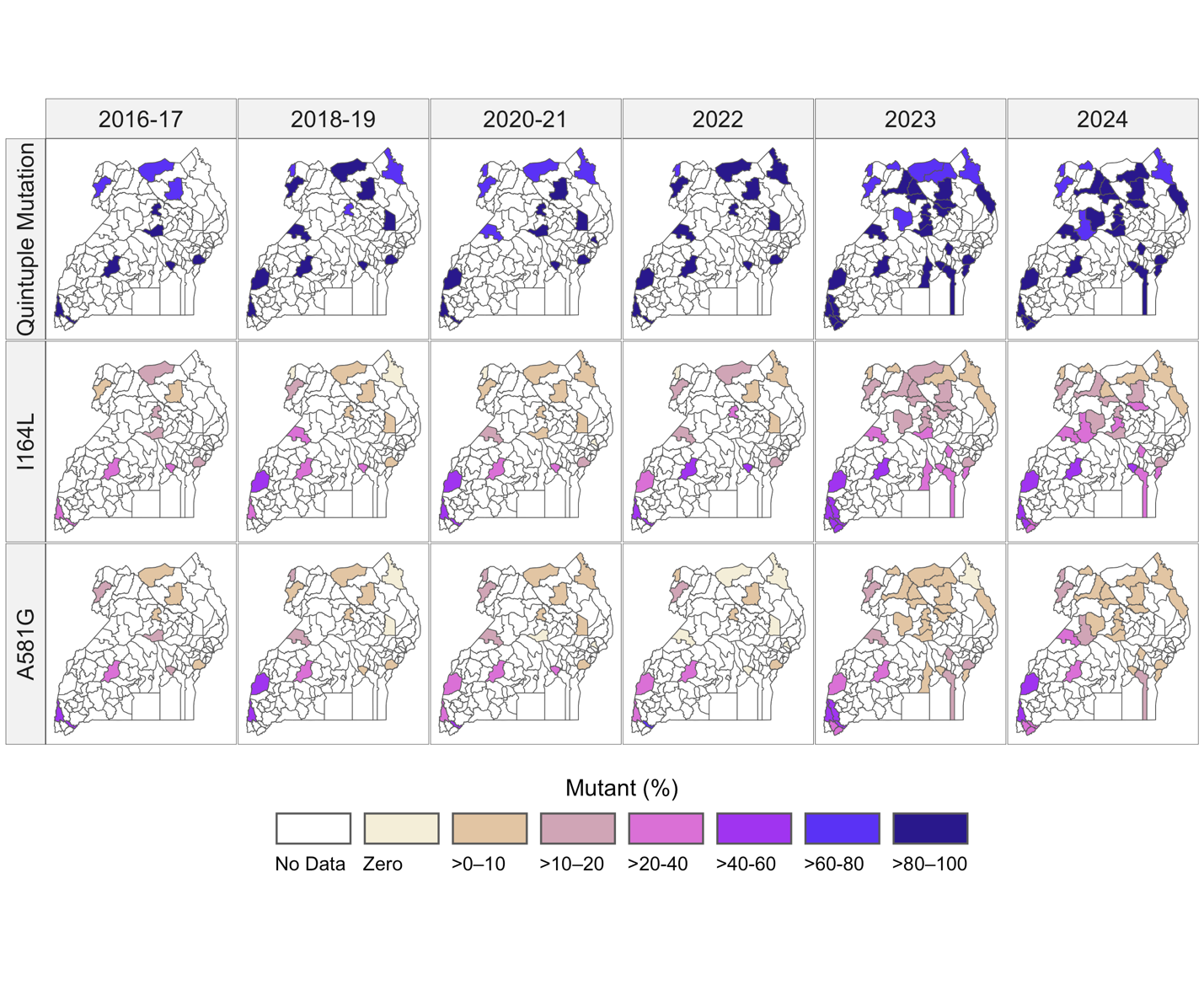

| Supplemental Table 1. Description of sample sites. | | | | | | | |
| --- | --- | --- | --- | --- | --- | --- | --- |
|  |  |  |  | Round 1 | | | |
|  |  |  |  | January - June 2023 | | | |
| Site | District | Sample collection strategy | MI1000 | N | Median age in years (IQR) | N samples collected from females (%) | Median parasite density by qPCR (IQR) |
| Aboke | Kole | biannual | 500 | 99 | 11 (6-15) | 75 (75.8%) | 5855.9 (786.8-30222.9) |
| Aduku | Kwania | biannual | 594 | 82 | 7 (4-15) | 51 (62.2%) | 23100.2 (7386.8-79645.2) |
| Alebtong | Alebtong | biannual | 136 | 86 | 11 (6-15) | 53 (61.6%) | 7177 (2711.9-44751.2) |
| Amolatar | Amolatar | biannual | 164 | 82 | 12 (5-24) | 50 (61%) | 4139.9 (831.3-23681.2) |
| Atiak | Amuru | biannual | 288 | 100 | 9 (4-13) | 69 (69%) | 18481.9 (3809.4-63619.5) |
| Awach | Gulu | biannual | 547 | 0 | - | - | - |
| Ayipe | Koboko | biannual | 794 | 0 | - | - | - |
| Bikurungu | Rukungiri | biannual | NA | 100 | 11 (6-15) | 55 (55%) | 6705.7 (3058.7-20190.4) |
| Budondo | Jinja | biannual | 195 | 77 | 9 (4-15) | 45 (58.4%) | 34407.3 (10281.1-92034.4) |
| Busitema | Busia | biannual | 1023 | 72 | 12 (5-18.5) | 42 (58.3%) | 6889.5 (391.2-39235.3) |
| Diima | Kiryandongo | biannual | 526 | 0 | - | - | - |
| Kamwezi | Rukiga | biannual | 107 | 93 | 13 (5-19) | 56 (60.2%) | 18450.8 (6808.8-45245.4) |
| Karambi | Kasese | biannual | 148 | 83 | 13 (10-17.5) | 36 (43.4%) | 15786.2 (4281.6-46800.4) |
| Kasambya | Mubende | biannual | 110 | 98 | 7 (3-15) | 55 (56.1%) | 18031.8 (4242.3-69924.2) |
| Kigandalo | Mayuge | biannual | 501 | 96 | 4 (2-9) | 53 (55.2%) | 2312.5 (191.5-12431.9) |
| Kigorobya | Hoima | biannual | 134 | 93 | 7 (3-12) | 50 (53.8%) | 28326.1 (6663.1-73300.5) |
| Kihihi | Kanungu | biannual | 98 | 100 | 12 (5-18) | 48 (48%) | 15978.1 (6373.2-59460.6) |
| Kiyunga | Mukono | biannual | 92 | 69 | 5 (2-16) | 42 (60.9%) | 17484.7 (2783.5-76955.7) |
| Kyatiri | Masindi | biannual | 179 | 0 | - | - | - |
| Lalogi | Omoro | biannual | 769 | 53 | 9.5 (3-15.5) | 41 (77.4%) | 2853.8 (692.3-20320.9) |
| Lokolia | Kaabong | biannual | 561 | 65 | 6.5 (2-15.2) | 34 (52.3%) | 1587.9 (199-16025.7) |
| Maziba | Kabale | year-round | 8 | 25 | 22 (9-36) | 12 (48%) | 16295.8 (4733.1-52604.1) |
| Metu | Moyo | biannual | 528 | 80 | 9 (5-13) | 52 (65%) | 18935.3 (4295.2-112944.7) |
| Muko | Rubanda | year-round | 10 | 81 | 23 (20-32) | 33 (40.7%) | 14078.3 (3728.7-46080.1) |
| Nadunget | Moroto | biannual | 391 | 56 | 4 (1-10) | 22 (39.3%) | 4212.7 (1045.9-18522.6) |
| Nagongera | Tororo | biannual | 243 | 91 | 8 (4-11) | 46 (50.5%) | 13842.7 (4270.7-49058.1) |
| Namokora | Kitgum | biannual | 537 | 100 | 14 (6.8-19) | 61 (61%) | 1853.2 (536.1-9640.7) |
| Nawaikoke | Kaliro | biannual | 345 | 94 | 3 (1-5.2) | 41 (43.6%) | 5690 (252.8-24590.5) |
| Opia | Arua | biannual | 349 | 93 | 13 (10-16) | 54 (58.1%) | 4710.7 (294-28600.3) |
| Orum | Otuke | biannual | 313 | 49 | 10 (4-13) | 28 (57.1%) | 19644.4 (1518-104272) |
| Padibe | Lamwo | biannual | 336 | 80 | 12 (7-15) | 45 (56.2%) | 6963.5 (887.2-33935.3) |
| Patongo | Agago | biannual | 406 | 97 | 9 (5-15) | 58 (59.8%) | 9976 (2865.8-36651.3) |

|  |  |  |  | Round 2 | | | |
| --- | --- | --- | --- | --- | --- | --- | --- |
|  |  |  |  | July - December 2023 | | | |
| Site | District | Sample collection strategy | MI1000 | N | Median age in years (IQR) | N samples collected from females (%) | Median parasite density by qPCR (IQR) |
| Aboke | Kole | biannual | 500 | 80 | 7 (2-13.2) | 50 (62.5%) | 36680.6 (7557.8-128850.8) |
| Aduku | Kwania | biannual | 594 | 82 | 9.5 (5-15) | 51 (62.2%) | 10350 (3715.2-48102.7) |
| Alebtong | Alebtong | biannual | 136 | 0 | - | - | - |
| Amolatar | Amolatar | biannual | 164 | 71 | 11 (7-14) | 43 (60.6%) | 5128.5 (2209.3-17377.1) |
| Atiak | Amuru | biannual | 288 | 99 | 7 (2-13.5) | 59 (59.6%) | 12685.1 (3686.5-49692.5) |
| Awach | Gulu | biannual | 547 | 98 | 9 (5-13) | 63 (64.3%) | 5578.8 (1822.2-14570.6) |
| Ayipe | Koboko | biannual | 794 | 71 | 6 (3-10) | 50 (70.4%) | 37824 (8296.7-92477) |
| Bikurungu | Rukungiri | biannual | NA | 0 | - | - | - |
| Budondo | Jinja | biannual | 195 | 67 | 8.5 (4-12.8) | 44 (65.7%) | 15582 (7142.6-42121.4) |
| Busitema | Busia | biannual | 1023 | 98 | 9 (2-13.8) | 62 (63.3%) | 9082.9 (941-38349.8) |
| Diima | Kiryandongo | biannual | 526 | 52 | 6 (3-10) | 28 (53.8%) | 14290.6 (6875.5-29704.5) |
| Kamwezi | Rukiga | biannual | 107 | 100 | 13.5 (9.8-21.2) | 57 (57%) | 17086.6 (5098.3-77230.2) |
| Karambi | Kasese | biannual | 148 | 70 | 13 (8.2-18) | 43 (61.4%) | 14089.1 (4288.3-36146.9) |
| Kasambya | Mubende | biannual | 110 | 93 | 10 (4-18) | 55 (59.1%) | 6429.8 (1445-28369.6) |
| Kigandalo | Mayuge | biannual | 501 | 99 | 4 (2-7) | 41 (41.4%) | 8749.4 (3106.7-32515.5) |
| Kigorobya | Hoima | biannual | 134 | 99 | 7 (3.5-13) | 54 (54.5%) | 25959.2 (7966.1-75910.1) |
| Kihihi | Kanungu | biannual | 98 | 98 | 11 (5-17) | 56 (57.1%) | 17065.7 (8678.6-81985.1) |
| Kiyunga | Mukono | biannual | 92 | 0 | - | - | - |
| Kyatiri | Masindi | biannual | 179 | 0 | - | - | - |
| Lalogi | Omoro | biannual | 769 | 44 | 8 (1-13.2) | 26 (59.1%) | 13494.9 (2449.4-44020) |
| Lokolia | Kaabong | biannual | 561 | 99 | 6 (2-14) | 69 (69.7%) | 15876.2 (2916-61499.5) |
| Maziba | Kabale | year-round | 8 | 27 | 21 (18-35) | 7 (25.9%) | 2455.1 (426.7-6515.4) |
| Metu | Moyo | biannual | 528 | 91 | 8 (3.5-13.5) | 50 (54.9%) | 22083.7 (5918.6-77744.6) |
| Muko | Rubanda | year-round | 10 | 34 | 20 (18-24) | 3 (8.8%) | 5181.2 (1648.9-10831.5) |
| Nadunget | Moroto | biannual | 391 | 100 | 2 (1-6.2) | 54 (54%) | 20083.1 (5615.8-60976.4) |
| Nagongera | Tororo | biannual | 243 | 94 | 9 (4-15) | 54 (57.4%) | 3787.4 (677.9-38413.1) |
| Namokora | Kitgum | biannual | 537 | 96 | 4 (2-10) | 50 (52.1%) | 21175.6 (5894.4-52464.3) |
| Nawaikoke | Kaliro | biannual | 345 | 65 | 3 (1-7) | 26 (40%) | 96492.4 (12600.9-1029934.5) |
| Opia | Arua | biannual | 349 | 77 | 12 (7-17) | 48 (62.3%) | 13537 (3939.8-72005.9) |
| Orum | Otuke | biannual | 313 | 76 | 8 (3-15) | 40 (52.6%) | 5376 (2490.4-10507.7) |
| Padibe | Lamwo | biannual | 336 | 99 | 12 (5-14.5) | 56 (56.6%) | 16473.4 (4930.1-42156.4) |
| Patongo | Agago | biannual | 406 | 100 | 12.5 (7-16) | 60 (60%) | 9345.3 (2705.8-25435.9) |

|  |  |  |  | Round 3 | | | |
| --- | --- | --- | --- | --- | --- | --- | --- |
|  |  |  |  | January - June 2024 | | | |
| Site | District | Sample collection strategy | MI1000 | N | Median age in years (IQR) | N samples collected from females (%) | Median parasite density by qPCR (IQR) |
| Aboke | Kole | biannual | 500 | 88 | 9.5 (3-13) | 52 (59.1%) | 9539.6 (2704.8-34150.3) |
| Aduku | Kwania | biannual | 594 | 72 | 12 (6.8-17) | 54 (75%) | 20052.2 (6462.5-59919.3) |
| Alebtong | Alebtong | biannual | 136 | 0 | - | - | - |
| Amolatar | Amolatar | biannual | 164 | Samples Excluded | | | |
| Atiak | Amuru | biannual | 288 | 51 | 10 (4-14) | 31 (60.8%) | 2705 (725.2-6744.9) |
| Awach | Gulu | biannual | 547 | 46 | 11 (4.2-13) | 23 (50%) | 15729.2 (3852.1-73146.4) |
| Ayipe | Koboko | biannual | 794 | 87 | 10 (3-15) | 56 (64.4%) | 3800 (1063.8-15802.6) |
| Bikurungu | Rukungiri | biannual | NA | 0 | - | - | - |
| Budondo | Jinja | biannual | 195 | 54 | 5 (2-8) | 24 (44.4%) | 17267.5 (3169-64868.7) |
| Busitema | Busia | biannual | 1023 | 59 | 10 (3-19) | 33 (55.9%) | 2691.3 (640.6-12964.2) |
| Diima | Kiryandongo | biannual | 526 | 100 | 8 (4-13.2) | 60 (60%) | 9107.3 (1371.4-37540.4) |
| Kamwezi | Rukiga | biannual | 107 | 164 | 15 (10-20) | 78 (47.6%) | 3720 (471.1-16614.9) |
| Karambi | Kasese | biannual | 148 | 38 | 13.5 (10.2-17.8) | 19 (50%) | 4862.4 (844.9-21196.5) |
| Kasambya | Mubende | biannual | 110 | 100 | 10 (4-14) | 59 (59%) | 12238.6 (2291.8-55987.7) |
| Kigandalo | Mayuge | biannual | 501 | 99 | 4 (1-11) | 57 (57.6%) | 24210.8 (3874.1-150408.2) |
| Kigorobya | Hoima | biannual | 134 | 100 | 8 (4-13) | 61 (61%) | 15196 (3692.2-80880.6) |
| Kihihi | Kanungu | biannual | 98 | 13 | 10 (3-15) | 4 (30.8%) | 2031 (524.1-11366) |
| Kiyunga | Mukono | biannual | 92 | 0 | - | - | - |
| Kyatiri | Masindi | biannual | 179 | 88 | 6 (3-10) | 42 (47.7%) | 14583.2 (4725.7-49916.7) |
| Lalogi | Omoro | biannual | 769 | 28 | 13.5 (7-20.2) | 22 (78.6%) | 36843.8 (9787.7-144054.4) |
| Lokolia | Kaabong | biannual | 561 | 54 | 13 (4-21.8) | 39 (72.2%) | 755.9 (274.8-4524) |
| Maziba | Kabale | year-round | 8 | 43 | 22 (11.5-30) | 26 (60.5%) | 9622.6 (3644.8-75983.9) |
| Metu | Moyo | biannual | 528 | 77 | 14 (11-16) | 43 (55.8%) | 905 (271.4-4004.9) |
| Muko | Rubanda | year-round | 10 | 52 | 24 (19-29.8) | 16 (30.8%) | 9786.6 (2314.9-52818.8) |
| Nadunget | Moroto | biannual | 391 | 99 | 7 (2-15) | 59 (59.6%) | 4637.3 (1191.8-43057.1) |
| Nagongera | Tororo | biannual | 243 | 96 | 6 (2-12.2) | 61 (63.5%) | 28932.4 (7418.4-74019.6) |
| Namokora | Kitgum | biannual | 537 | 93 | 8 (3-13) | 53 (57%) | 11324 (4293.4-60901.1) |
| Nawaikoke | Kaliro | biannual | 345 | 78 | 4 (1-9.8) | 49 (62.8%) | 20646.3 (2323-92722.4) |
| Opia | Arua | biannual | 349 | 33 | 13 (10-15) | 21 (63.6%) | 764.7 (404-5781.2) |
| Orum | Otuke | biannual | 313 | 46 | 12 (6-16) | 29 (63%) | 3656.6 (1480.1-26480.4) |
| Padibe | Lamwo | biannual | 336 | Samples Excluded | | | |
| Patongo | Agago | biannual | 406 | 100 | 14 (10-16.2) | 69 (69%) | 4188.2 (1197.4-20952) |

|  |  |  |  | Round 4 | | | |
| --- | --- | --- | --- | --- | --- | --- | --- |
|  |  |  |  | July - December 2024 | | | |
| Site | District | Sample collection strategy | MI1000 | N | Median age in years; (IQR) | N samples collected from females (%) | Median parasite density by qPCR (IQR) |
| Aboke | Kole | biannual | 500 | 59 | 7 (4-19) | 37 (62.7%) | 17120 (5202.6-59052.9) |
| Aduku | Kwania | biannual | 594 | Samples Excluded | | | |
| Alebtong | Alebtong | biannual | 136 | 0 | - | - | - |
| Amolatar | Amolatar | biannual | 164 | 66 | 13 (5-19) | 52 (78.8%) | 11261.9 (3628.4-37249.3) |
| Atiak | Amuru | biannual | 288 | 100 | 7 (3-12.2) | 61 (61%) | 17143.5 (4842.6-64031.1) |
| Awach | Gulu | biannual | 547 | 74 | 10 (2.2-14) | 41 (55.4%) | 11945 (3092.1-65485.4) |
| Ayipe | Koboko | biannual | 794 | 56 | 11.5 (8.8-20) | 34 (60.7%) | 4731.6 (978.3-19654.7) |
| Bikurungu | Rukungiri | biannual | NA | 0 | - | - | - |
| Budondo | Jinja | biannual | 195 | 69 | 8 (3-17) | 42 (60.9%) | 21831.5 (2250.9-160827.4) |
| Busitema | Busia | biannual | 1023 | 42 | 18 (9.2-26.2) | 29 (69%) | 2426.1 (925.6-17731.5) |
| Diima | Kiryandongo | biannual | 526 | 100 | 8 (3-13.2) | 62 (62%) | 15607.5 (2952.3-65077.9) |
| Kamwezi | Rukiga | biannual | 107 | 32 | 18.5 (12.5-25) | 16 (50%) | 8885.6 (2339.7-30025) |
| Karambi | Kasese | biannual | 148 | 43 | 16 (9-23.5) | 31 (72.1%) | 19744.2 (624.5-65067.2) |
| Kasambya | Mubende | biannual | 110 | 86 | 11.5 (5-16.8) | 48 (55.8%) | 12262.1 (1876.8-47988) |
| Kigandalo | Mayuge | biannual | 501 | 98 | 4 (1-7) | 55 (56.1%) | 18308.2 (4382.6-49944.8) |
| Kigorobya | Hoima | biannual | 134 | 98 | 9.5 (5-12) | 50 (51%) | 50069 (11041.2-241026.5) |
| Kihihi | Kanungu | biannual | 98 | 29 | 12 (7-17) | 16 (55.2%) | 2693 (638.1-33435.5) |
| Kiyunga | Mukono | biannual | 92 | 0 | - | - | - |
| Kyatiri | Masindi | biannual | 179 | 78 | 5.5 (2-9.8) | 45 (57.7%) | 25280.4 (5804.8-122095.9) |
| Lalogi | Omoro | biannual | 769 | Samples Excluded | | | |
| Lokolia | Kaabong | biannual | 561 | 57 | 4 (3-9) | 23 (40.4%) | 2213.3 (434.8-22849.1) |
| Maziba | Kabale | year-round | 8 | 19 | 21 (16.5-26) | 4 (21.1%) | 11925.4 (7549-20344) |
| Metu | Moyo | biannual | 528 | 28 | 12.5 (9-14.5) | 14 (50%) | 7289.4 (1924.4-15139.2) |
| Muko | Rubanda | year-round | 10 | 20 | 29 (21-32) | 7 (35%) | 4602.3 (1229.8-16948.9) |
| Nadunget | Moroto | biannual | 391 | 100 | 3 (2-5.2) | 51 (51%) | 57373.7 (7590.6-456790.3) |
| Nagongera | Tororo | biannual | 243 | 21 | 7 (2-10) | 11 (52.4%) | 3989.5 (993-22981.2) |
| Namokora | Kitgum | biannual | 537 | 100 | 4 (1.8-10) | 52 (52%) | 53129.8 (13984-170131) |
| Nawaikoke | Kaliro | biannual | 345 | 96 | 5 (2-10) | 45 (46.9%) | 4111.9 (391.6-51659) |
| Opia | Arua | biannual | 349 | 36 | 10.5 (7.8-14.2) | 27 (75%) | 131.3 (61.5-1738.7) |
| Orum | Otuke | biannual | 313 | 97 | 9 (5-14) | 59 (60.8%) | 31157.8 (6998.9-120623.2) |
| Padibe | Lamwo | biannual | 336 | Samples Excluded | | | |
| Patongo | Agago | biannual | 406 | 100 | 11 (5-15) | 70 (70%) | 17649.9 (4436.8-75201.2) |

**Supplemental Table 2. Prevalence and frequency of markers of antimalarial resistance by collection.** Available at: 10.5281/zenodo.18854846

**Supplemental Table 3. Tabulation of samples with increased copy number by year. Also available at** 10.5281/zenodo.18854846

| Sample Site | Year | MDR1 | PM |
| --- | --- | --- | --- |
| Aboke | 2023 | 0 / 138 (0.0%) | 0 / 138 (0.0%) |
| Aboke | 2024 | 0 / 148 (0.0%) | 0 / 148 (0.0%) |
| Aduku | 2023 | 1 / 163 (0.6%) | 0 / 163 (0.0%) |
| Aduku | 2024 | 0 / 77 (0.0%) | 0 / 77 (0.0%) |
| Alebtong | 2023 | 0 / 88 (0.0%) | 0 / 88 (0.0%) |
| Amolatar | 2023 | 0 / 136 (0.0%) | 0 / 136 (0.0%) |
| Amolatar | 2024 | 0 / 74 (0.0%) | 0 / 74 (0.0%) |
| Atiak | 2023 | 1 / 176 (0.6%) | 0 / 176 (0.0%) |
| Atiak | 2024 | 0 / 137 (0.0%) | 0 / 137 (0.0%) |
| Awach | 2023 | 0 / 97 (0.0%) | 0 / 97 (0.0%) |
| Awach | 2024 | 0 / 138 (0.0%) | 0 / 138 (0.0%) |
| Ayipe | 2023 | 0 / 71 (0.0%) | 0 / 71 (0.0%) |
| Ayipe | 2024 | 0 / 147 (0.0%) | 0 / 147 (0.0%) |
| Bikurungu | 2023 | 0 / 96 (0.0%) | 0 / 96 (0.0%) |
| Budondo | 2023 | 1 / 104 (1.0%) | 0 / 104 (0.0%) |
| Budondo | 2024 | 0 / 136 (0.0%) | 0 / 136 (0.0%) |
| Busitema | 2023 | 1 / 128 (0.8%) | 0 / 128 (0.0%) |
| Busitema | 2024 | 0 / 99 (0.0%) | 0 / 99 (0.0%) |
| Diima | 2023 | 0 / 51 (0.0%) | 0 / 51 (0.0%) |
| Diima | 2024 | 1 / 191 (0.5%) | 0 / 191 (0.0%) |
| Kamwezi | 2023 | 0 / 172 (0.0%) | 0 / 172 (0.0%) |
| Kamwezi | 2024 | 4 / 182 (2.2%) | 0 / 182 (0.0%) |
| Karambi | 2023 | 1 / 151 (0.7%) | 0 / 151 (0.0%) |
| Karambi | 2024 | 0 / 78 (0.0%) | 0 / 78 (0.0%) |
| Kasambya | 2023 | 1 / 175 (0.6%) | 0 / 175 (0.0%) |
| Kasambya | 2024 | 3 / 141 (2.1%) | 0 / 141 (0.0%) |
| Kigandalo | 2023 | 0 / 170 (0.0%) | 0 / 170 (0.0%) |
| Kigandalo | 2024 | 0 / 186 (0.0%) | 0 / 186 (0.0%) |
| Kigorobya | 2023 | 0 / 150 (0.0%) | 0 / 150 (0.0%) |
| Kigorobya | 2024 | 0 / 186 (0.0%) | 0 / 186 (0.0%) |
| Kihihi | 2023 | 0 / 174 (0.0%) | 0 / 174 (0.0%) |
| Kihihi | 2024 | 0 / 41 (0.0%) | 0 / 41 (0.0%) |
| Kiyunga | 2023 | 0 / 72 (0.0%) | 0 / 72 (0.0%) |
| Kyatiri | 2024 | 0 / 142 (0.0%) | 0 / 142 (0.0%) |
| Lalogi | 2023 | 0 / 127 (0.0%) | 0 / 127 (0.0%) |
| Lalogi | 2024 | 0 / 47 (0.0%) | 1 / 47 (2.1%) |
| Lokolia | 2023 | 0 / 146 (0.0%) | 0 / 146 (0.0%) |
| Lokolia | 2024 | 0 / 107 (0.0%) | 0 / 107 (0.0%) |
| Maziba | 2023 | 0 / 50 (0.0%) | 0 / 50 (0.0%) |
| Maziba | 2024 | 0 / 60 (0.0%) | 0 / 60 (0.0%) |
| Metu | 2023 | 0 / 157 (0.0%) | 0 / 157 (0.0%) |
| Metu | 2024 | 0 / 97 (0.0%) | 0 / 97 (0.0%) |
| Muko | 2023 | 0 / 100 (0.0%) | 1 / 100 (1.0%) |
| Muko | 2024 | 0 / 69 (0.0%) | 0 / 69 (0.0%) |
| Nadunget | 2023 | 0 / 150 (0.0%) | 0 / 150 (0.0%) |
| Nadunget | 2024 | 0 / 194 (0.0%) | 0 / 194 (0.0%) |
| Nagongera | 2023 | 0 / 100 (0.0%) | 0 / 100 (0.0%) |
| Nagongera | 2024 | 0 / 71 (0.0%) | 1 / 71 (1.4%) |
| Namokora | 2023 | 0 / 162 (0.0%) | 0 / 162 (0.0%) |
| Namokora | 2024 | 0 / 194 (0.0%) | 0 / 194 (0.0%) |
| Nawaikoke | 2023 | 0 / 136 (0.0%) | 0 / 136 (0.0%) |
| Nawaikoke | 2024 | 1 / 165 (0.6%) | 1 / 165 (0.6%) |
| Opia | 2023 | 0 / 159 (0.0%) | 0 / 159 (0.0%) |
| Opia | 2024 | 0 / 69 (0.0%) | 0 / 69 (0.0%) |
| Orum | 2023 | 0 / 122 (0.0%) | 0 / 122 (0.0%) |
| Orum | 2024 | 0 / 142 (0.0%) | 0 / 142 (0.0%) |
| Padibe | 2023 | 0 / 168 (0.0%) | 0 / 168 (0.0%) |
| Patongo | 2023 | 1 / 110 (0.9%) | 0 / 110 (0.0%) |
| Patongo | 2024 | 0 / 197 (0.0%) | 0 / 197 (0.0%) |

**Supplemental Table 4. Prevalence and frequency of DHFR and DHPS haplotypes by collection.** Available at: 10.5281/zenodo.18854846
